# The expression levels of *NOS2, HMOX1* and *VEGFC* in cumulus cells are markers of oocyte maturation and fertilization rate

**DOI:** 10.1101/2022.07.22.22277925

**Authors:** Montserrat Barragán, David Cornet-Bartolomé, Natalia Molina, Rita Vassena

## Abstract

**Purpose:** We aim to study whether the expression of genes related to hypoxia and ageing in cumulus cells (CC) is associated to reproductive results in *in vitro* fertilization cycles.

**Methods:** Cumulus cells (CC) were collected after ovum pick-up (OPU) from 94 women recruited undergoing controlled ovarian stimulation from 2018 to 2021; total RNA was extracted and reverse transcribed to cDNA using random hexamers. 14 genes affected by ageing in humans, and expressed in CC, were selected for testing. Expression levels of these genes were detected by qPCR and normalized against *TATA-box binding protein (TBP) in CC*. Expression levels were plotted against woman age, AFC, MII rate at OPU (MR), and fertilization rates (FR) and non-parametric Spearman’s correlation was applied.

**Results:** From the initial list of 423 candidate genes, we tested 14 genes related to hypoxia response via HIF1alpha activation, oxidative stress response, and angiogenic response. The expression of *CLU, NOS2* and *TXNIP* had a positive correlation with age (rs=0.25, p=0.014; rs=0.31, p=0.0027; and rs=0.24, p=0.03; respectively). Additionally, *NOS2* and *HMOX1* expression correlated positively with the retrieval of immature oocytes (rs = 0.23228, p = 0.03242; rs = 0.38827, p = 0.01212; respectively). Moreover, *VEGFC* levels decreased overall with increasing fertilization rate, independently of age (rs=-0.29, p=0.026).

**Conclusion:** We found that the fertilization potential of a cohort of oocytes is related to the ability of CC to respond to oxidative stress and hypoxia, pointing at NOS2, HMOX1 and VEGFC expression as markers for oocyte maturation and fertilization success.

## INTRODUCTION

Aging is a risk factor for the appearance of several diseases, including infertility [1,2].

Advanced maternal age (AMA) is defined as childbearing past 35 years of age. In resources rich countries, the average age at first birth is on the rise and increasingly the process is spreading to middle- and low-income countries [3-6].

Women are born with a limited oocyte pool in their ovaries constituted in the foetus at 18–22 weeks post-conception [7]. This pool gradually decreases through the reproductive lifespan, becoming close to zero at menopause [7, 8-9]. The loss of fertility over time is mostly caused by a decrease in ovarian reserve [10], increased chromosomal abnormalities in gametes and embryos, decreased oocyte developmental competence [11-12], and pregnancy complications [13-14].

Oocyte developmental competence is defined as the ability of the oocyte to sustain embryonic development at least until the embryonic genome activation (EGA) stage [15-16]. To acquire its developmental competence, the oocyte grows and mature through a continuous crosstalk between with the somatic follicular cells (cumulus cells, CC) that surround it. Throughout a woman reproductive life, cumulus cells (CC) protect the dormant oocyte from damage, act as sensors of the follicular microenvironment and as gatekeepers of the oocyte developmental potential.

Follicles, CC and oocytes differentially express specific pattern of genes as they age, affecting oocyte maturation and, possibly, altering the developmental competence of the maturing oocyte. Several studies have identified non-invasive markers to predict oocyte quality, embryo development and pregnancy outcomes by analyzing the transcriptome of human CCs [17-21], including comparisons between CC from young and older women [22-24], Recently, alterations in CC response to the follicular microenvironment have been described in AMA [25-26]. One such pathway is the hypoxia-dependent response, which controls the cell metabolic and oxidative state. With age, hypoxia tolerance decreases in the whole organism [27], however whether ovarian tissue could age earlier than other tissues is still a matter of study.

Here, we investigate whether expression levels of aging-related genes in CC could provide insights into the competence of the oocyte cohort in relation to maternal age.

## MATERIALS AND METHODS

### Study population

We included 94 women from March 2018 to March 2021, that underwent ICSI/IVF cycles using their own oocytes and sperm donor (IVF, n=53) or participating in an oocyte donation program (DON, n=41). Exclusion criteria were: a diagnosis of premature ovarian failure (POF) or polycystic ovary syndrome (PCOS).

### Ovarian stimulation

Controlled ovarian stimulation was induced with either Follitropin alpha (Gonal®, Merck-Serono, Spain) or highly purified hMG (Menopur®, Ferring, Spain), with daily injections of 150-300 IU [28]. Pituitary suppression was performed with a GnRH antagonist (0.25 mg of Cetrorelix acetate, Cetrotide®, Merck Serono, Spain) administered daily from Day 6 of stimulation [29].

When a minimum of 3 follicles of >18 mm of diameter, and at least 5 follicles of ≥ 16 mm, developed on both ovaries, final oocyte maturation and ovulation was triggered with either 0.3 mg of GnRH agonist (Decapeptyl®, Ipsen Pharma S.A., Spain) or 250 μg hCG (Ovitrelle®, Merck, Germany), in DON and IVF women, respectively.

### Cumulus cells collection and processing

Ovum pick-up (OPU) was performed 36 hours after triggering. Cumulus oocyte complexes (COCs) were collected in buffered medium (G-MOPS® PLUS, Vitrolife, Göteborg), containing Human Serum Albumin (HSA; Vitrolife) and incubated at 37°C in 6%CO_2_ and 95% relative humidity for thirty minutes before CC removal by exposure to 80 IU/mL of hyaluronidase (HYASE-10x®, Vitrolife). CCs were mechanically isolated by gentle pipetting in drops of buffered medium (G-MOPS^®^ PLUS, Vitrolife) using capillaries of progressively smaller diameter (Flexi-Pet^®^; Cook Medicals, Bloomington, IN). Once isolated, pooled CCs were collected from each woman and resuspended in 350 μl of RNA lysis buffer (RTL, RNAeasy Mini Kit, QIAGEN, Germany), stored at 4ºC and processed the same day for total RNA extraction.

### Selection of aging-related genes

To select aging-related patterns in CCs, we first obtained a list of established and experimentally verified aging-related human genes (GenAge [30]), and cross-referenced it with the published human CC transcriptome [19] using Venny algorithm [31]. The complete list of these genes can be found in Supplementary Table 1. A further GO enrichment [32] and STRING [33] analyses were applied to identify the set of genes to be analyzed.

**Table 1.**
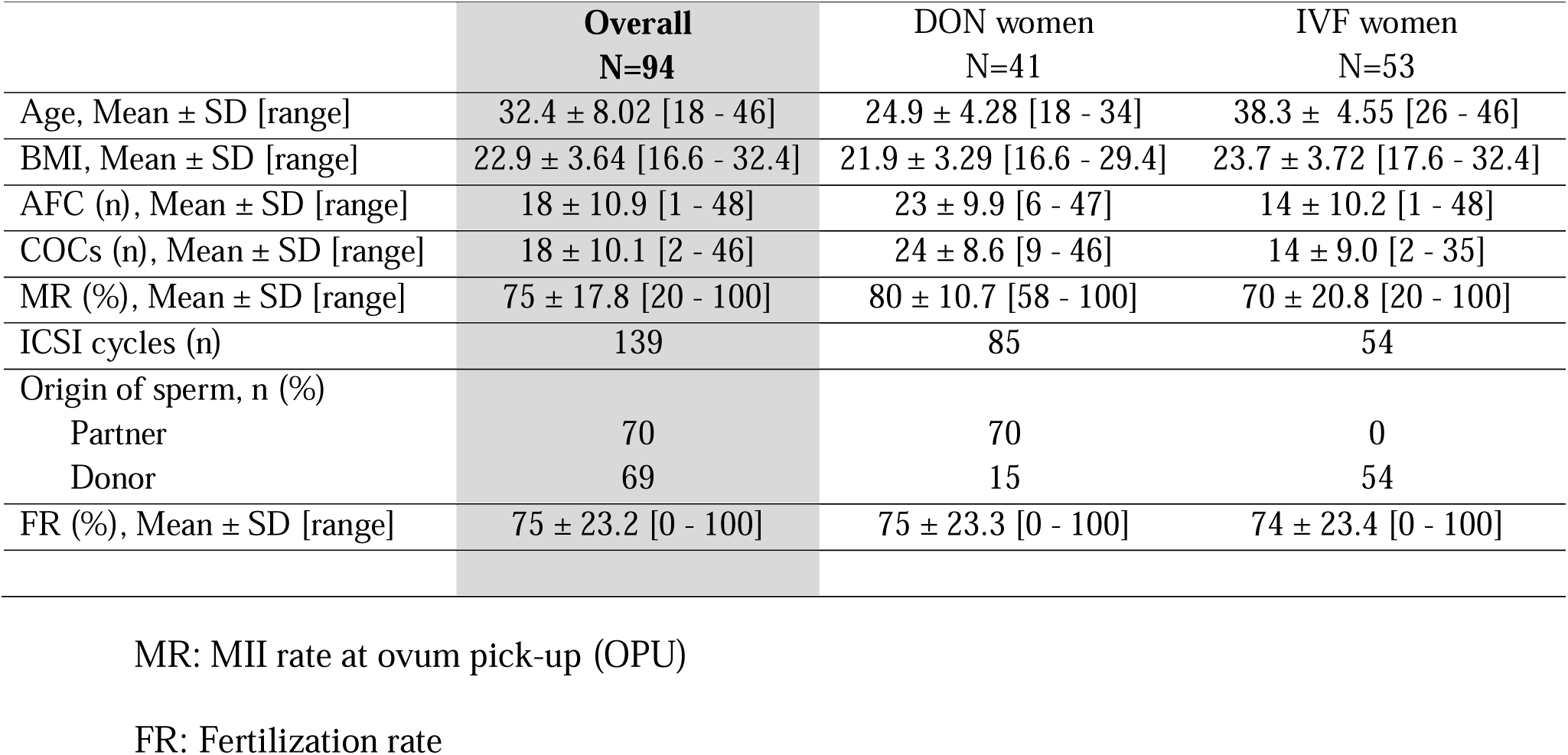
Demographic characteristics.

### Total RNA extraction and cDNA synthesis

Total RNA was isolated with RNeasy Mini Kit (Qiagen) according to the manufacturer’s protocol. Total RNA was quantified using Quawell3000 by measurement of absorbance at λ=260nm and stored at -80ºC. For cDNA synthesis, equal amounts of total RNA (from 800 ng to 1 μg) from each sample were reverse transcribed to cDNA in a CFX96 Real-Time PCR system (Bio-Rad) with the SuperScript™ IV (SSIV) First-Strand Synthesis Kit (Invitrogen) using random hexamers and following the manufacturer’s protocol. Briefly, cDNA synthesis was produced as follow: primer annealing was performed for 5 minutes at 65ºC (50 ng random hexamers, 10 nmol dNTP mix, and up to 5 μg total RNA), after a short incubation (2 minutes) at 4ºC a mixture of RT buffer, 100 nmol DTT, 40 U RNAse inhibitor and 200 U SSIV-RT was added and reverse transcription was performed for 10 min at 25 °C, 10 min at 55 °C, and 10 min at 80 °C. Synthetized cDNA was stored at -20 °C until use.

### qPCR analysis

Each qPCR reaction included 5ng of cDNA, 10μl 2x SsoAdvanced Universal SYBR Green Supermix (BioRad, Hercules CA, USA), and 10 pmol of each primer, in a 20μl final reaction volume. The program used for each qPCR run consisted of an initial denaturalization step of 30s at 95ºC, and 40 cycles of 95ºC for 5s and 60ºC for 30s (data collection point) followed by a melting curve analysis with data collected during each cycle at the 60 °C extension step with CFX Manager Software v3.0 (BioRad). All qPCRs were carried out in triplicate, including no-template controls, in 96-well plates using the CFX96 system (BioRad). Threshold and baselines were automatically determined and the specificity of amplicons was confirmed by melt curve analysis and Sanger gene sequencing.

Expression levels of selected genes, aging-(*AMH, ANXA5, ATP5G3, FGF2, LYZ*) and hypoxia-related genes (*CLU, FABP3, HMOX1, HIF-1A, NOS2, NOS3, TGFBR3, TXNIP, VEGFC*), were analyzed by qPCR first for their presence in cumulus cells and then for differential expression levels.

To determine the best normalization set of genes, four different algorithms (geNorm (https://genorm.cmgg.be), NormFinder (https://moma.dk/normfinder-software), BestKeeper (https://www.gene-quantification.de/bestkeeper.html), and the comparative Ct method (https://heartcure.com.au)) were applied to qPCR data over 9 putative reference genes (HK) (e.g., *ACTB, GAPDH, GUSB, RPLP0, SDHA, TBP, UBC, YWHAZ, 18S*) on 12 additional representative CC samples (mean age 32 years old, SD= 8.2, range [20-44]; mean AFC 23.3, SD= 9.58, range [11-42]). *TBP* was selected as the best normalizer gene in our sample set.

The corresponding normalization factor (Ref) was then used to correct the relative gene expression values: (ΔCq = [Cq (gene A) □ Cq (Ref)]). The following formula was applied to do ΔCq analysis: normalized target gene expression level = 2^-(ΔCq)^. Primer sequences and efficiencies are specified in Supplementary Table 2.

### Statistical analysis

The relative expression levels of studied genes were plotted against woman age, AFC, MII rate (MR) at OPU and fertilization rate (FR); and non-parametric analysis (Spearman’s rho, r_s_) was conducted to evaluate their correlation. P-value (2-sides) <0.05 was considered statistically significant.

## RESULTS

### The efficiency of stimulation was not affected by women age

Demographic and cycle characteristics of the participants are presented in Table 1.

The analysis of correlation between woman’s age and AFC (r_s_ = -0.531; p = 3.69E-08) confirmed the expected decrease in ovarian reserve with age and a negative trend was observed for the proportion of MII oocytes obtained compared to the number of COCs retrieved (MR) (r_s_ =-0.197; p = 0.0571).

However, the efficiency of stimulation, i.e., the number of COCs retrieved compared to AFC, was not affected by age (*r*_*s*_ = 0.12631, *p* = 0.22509). When fertilization rates were drawn against age, we did not find a significant linear trend with age (r_s_ = -0.1; p = 0.39).

### Selection of gene-set

From the 13,018 genes expressed in cumulus cells (independently of oocyte maturation), 423 have been annotated as aging-related genes in GenAge database (Figure 1a). GO analysis revealed metabolic and response to stimulus as the major enriched pathways, suggesting that expression of some of these genes can be correlated with cohort reproductive outcomes in relation to women’s age (Figure 1b). Response to stimulus highlighted, as expected, signaling pathways in response to FSH and gonadotropin receptors, angiogenesis, oxidative stress and hypoxia response via HIF1alpha activation. The last three pathways caught our attention to select the panel of 14 genes to be analyzed in cumulus cells from women of different ages (Figure 1c): *HIF1A, NOS2, NOS3*, and *HMOX1* for hypoxia response via HIF1alpha activation; *LYZ, TXNIP, CLU, FABP3*, and *ATP5G3* for oxidative stress response; *FGF2, TGFBR3, ANXA5*, and *VEGFC* for angiogenic response. In addition, we decided to include *AMH* as a possible marker for follicular aging process, as previously suggested by Kedem and colleagues [34].

**FIG 1.**
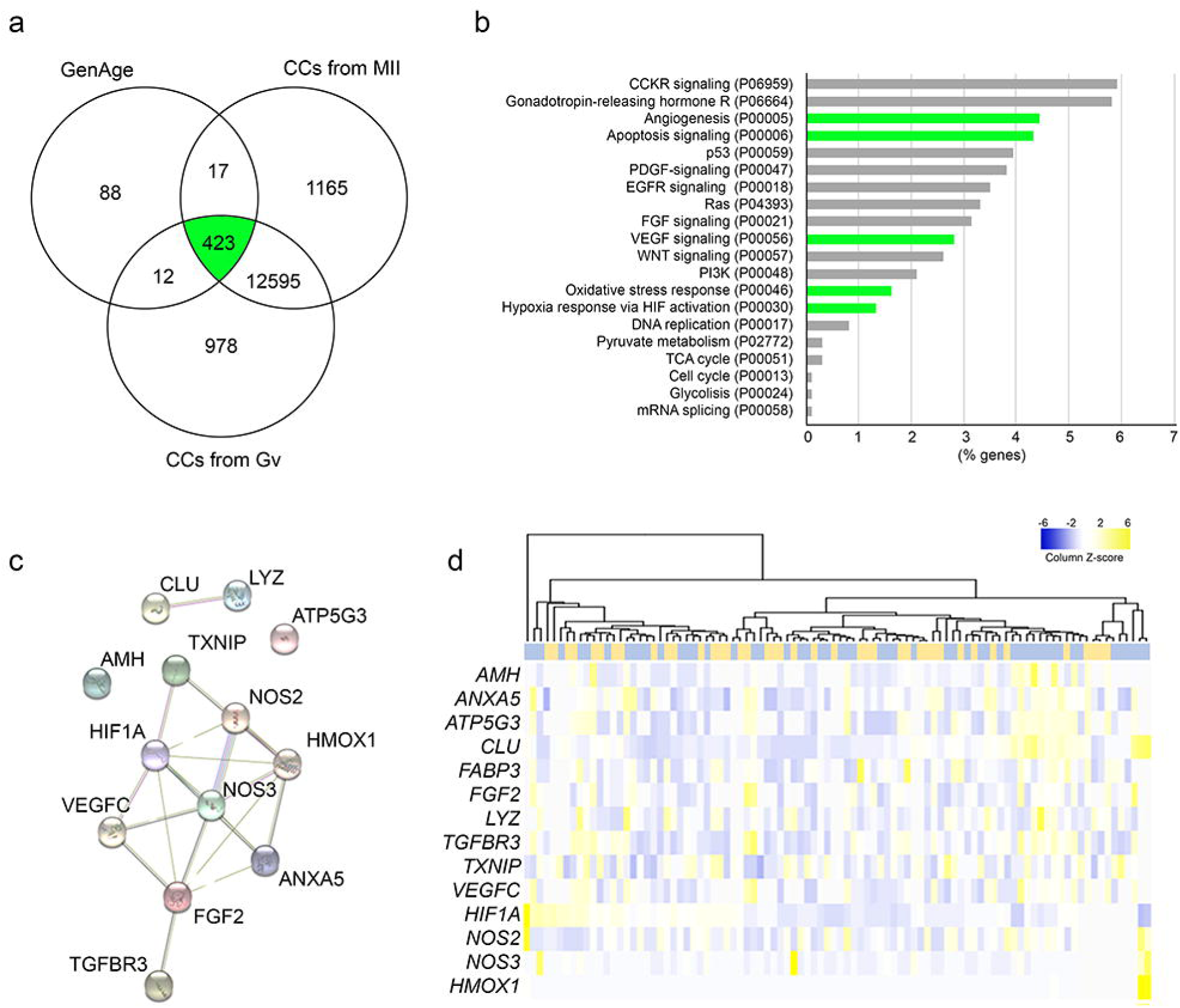
Selection of gene-set. **(a)** Venn diagrammatic representation of the cross-referenced genes found in “CCs from GV” *vs* “CCs from MII” *vs* “GeneAge”. **(b)** Top enriched pathways (GO) identified for the 423 aging-related genes that are expressed in CCs. **(c)** STRING analysis shows known interaction between the 14 selected genes. **(d)** Heat map for the expression of selected genes when CCs obtained from women (yellow: DON; blue: IVF) were compared and hierarchical clustering to analyze similarities between individualized samples. The colour in the heat map (blue to yellow scale) corresponds to relative transcript abundance

### Hypoxic adaptation in CCs as a marker of the oocyte cohort quality

Normalized expression levels of *AMH* were determined in CCs from women of different ages. We showed no correlation with any of the variables analyzed (age, AFC, obtained MII rate, or fertilization rate) (r_s_ <│0.25│, p>0.05).

Further, we analyzed the relative expression levels of all selected genes, showing different expression levels among CC (Supplementary Figure 1), while no clear clustering was observed on a heatmap (Figure 1d).

Further analysis of gene expression of selected genes presented positive correlation with age for three of them: *CLU* (a secreted chaperone involved in cell death) and two HIF1A-regulated genes, *NOS2* (an inducible and calcium-independent isoform of nitric oxide synthase) and *TXNIP* (a tumor suppressor gene involved in redox regulation), correlated positively with age (rs=0.25, p=0.014; rs=0.31, p=0.0027; and rs=0.24, p=0.03; respectively). When analyzing expression levels of aging markers (*LYZ, FGF2, ATP5G3, ANXA5*) in CCs, none of them correlate with age, AFC, or obtained MII rate (r_s_ <│0.1│, p>0.05).

Although abundant (Supplementary Figure 1), the expression of *HIF1A*, the master transcriptional regulator of cellular and developmental response to hypoxia, did not correlate with age, AFC, MII rate, or fertilization rate (r_s_ <│0.25│, p>0.05).

To further analyze HIF1A-dependent response, we first checked *NOS2* (an inducible and calcium-independent isoform of nitric oxide synthase) and *HMOX1* gene expression in CCs cohort in relation to clinical history and cycle outcomes, including fertilization rates. Overall, *NOS2* expression did not show correlation with AFC, obtained MII rate, or fertilization rate. However, *NOS2* expression showed a positive correlation with the presence of immature oocytes overall (rs = 0.23228, p = 0.03242) (Figure 2a), being relevant in very young (<23 years, rs = 0.62851, p = 0.01607) and oldest women (>40 years; rs = 0.51671, p = 0.03369).

**FIG 2.**
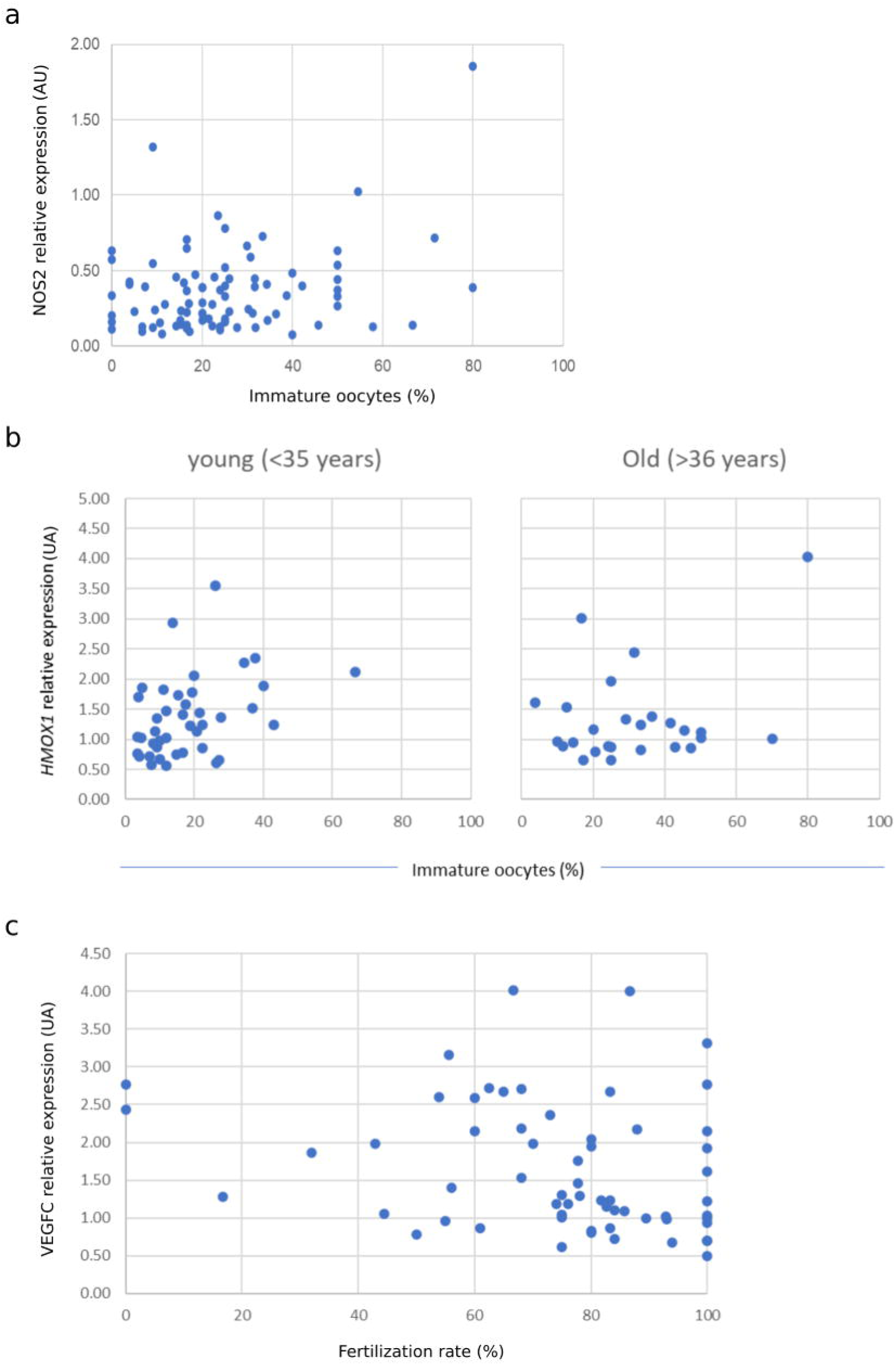
Correlation between gene expression levels and cycle outcomes. Relative abundance of gene expression levels was determined by real-time qPCR for **(a)** *NOS 2* plotted against overall immature rate (%) after OPU; **(b)** *HMOX1* plotted against immature rate (%) after OPU in young (<35 years) and AMA (>36 years). **(c)** *VEGFC* plotted against overall Fertilization rate (%)

In case of *HMOX1* expression, no correlation was observed with AFC, obtained MII rate, or fertilization rate; but a positive correlation was observed in young women (<35 years) with the presence of immature oocytes (rs = 0.38827, p (2-tailed) = 0.01212), and basal levels of *HMOX1* expression were observed in AMA women (>36 years) independently of the presence of immature oocytes in the COC cohort (Figure 2b).

To investigate further how hypoxic adaptation is reflected in cohort’s oocyte quality we measured the association between CCs gene expression and fertilization rate and we found that *VEGFC* levels decrease overall with increasing fertilization rate, independently of age (rs=-0.29, p=0.026) (Figure 2c).

## DISCUSSION

The overall lack of correlation between the gene expression signature in aged ovarian tissues and other aged human tissues suggests that ovaries at the end of their reproductive lifespan are not prematurely aged. Our results corroborate that the ovary seems to be the first organ to age, affecting general health outcomes [35].

Gene expression levels of *AMH* have been suggested to exert intrafollicular functions during the final stages of folliculogenesis, from antral to pre-ovulatory follicles [36]. We did not observe significant correlation between *AMH* expression and age or MII rate, contrary to the findings of Kedem and colleagues, where higher expression levels of *AMH* in CC were found in pre-ovulatory follicles containing immature oocytes (GV and MI oocytes) than those containing mature oocytes (MII) [37] and in CC from old (>40 years) women compared to young (<35 years) ones [34].

The indicated studies analysed CCs from individual COCs, thus we hypothesize that our analysis on cohort CCs cohort may mask individual differences among follicles.

To identify specific markers of reproductive aging, we focused on hypoxia, angiogenesis and redox imbalance adaptation, in agreement with a previous RNAseq study [22] pointing at genes involved in hypoxia stress response.

*HIF1A* is the subunit alpha of HIF-1 transcription factor, which acts as a master regulator of cellular response to hypoxia by activating the transcription of angiogenesis, apoptosis, and redox imbalance adaptation related genes (reviewed in [38]) and has been recently associated with steroidogenic activity regulation and oocyte developmental rates during *in vitro* maturation (IVM) in bovine [39].

We found sustained *HIF1A* gene expression in human CCs, in concordance with previous findings in *in vitro* culture of luteinised human granulosa cells [40]. However, no significant correlation or tendency has been observed with age, AFC, MR or FR.

Although mRNA levels seem not to be correlated with reproductive aging, the functional response could be. Thus, we decided to analyze canonical genes carrying hypoxia response elements (HRE) such as *NOS2* or *HMOX1*. An increased expression of *NOS2* and *HMOX1* at mRNA and protein level in CCs of non-fertilized oocytes compared to fertilized ones has been reported [41]. However, these results were obtained from COCs that contained mature oocytes. As results were not controlled for male factor or cohort COCs, a clear relation between *NOS2* and *HMOX1* gene expression and fertilization ability could not be stablished.

We aimed to test *NOS2* and *HMOX1* gene expression in CCs cohort in relation to age, clinical history and cycle outcomes, including fertilization rates. Our results in young woman (<35 years) showed increased expression of *HMOX1* in COCs cohorts containing more immature oocytes, suggesting anti-oxidant, anti-inflammatory and anti-apoptotic roles of *HMOX1* [42] during oocyte maturation in young women. Interestingly, basal levels of *HMOX1* expression were observed independently of the number of immature oocytes obtained at OPU in old women (>36 years), suggesting that this pathway is altered in CCs from AMA women.

In addition, we found that *VEGFC* expression negatively correlated with fertilization rates. *VEGFC* encodes a member of the vascular endothelial growth factor family, promotes angiogenesis and modulates blood vessels permeability. Follicular angiogenesis is a crucial event during follicular recruitment, maturation, ovulation, and the establishment of a corpus luteum [43]. Genes of the VEGF family are expressed *in vivo* in several cell types of the female reproductive system, including theca, cumulus, and granulosa cells where *VEGF* gene expression is hormonally regulated [44-45]. This last work showed higher protein levels of VEGF in CCs matched with non-fertilized oocytes after conventional IVF, suggesting a role for oocyte-cumulus complex hypoxia. However, so far, no study has addressed the potential use of *VEGF* expression as a marker for fertilization rate after ICSI in a cohort after ovarian stimulation.

This study suggests that, in physiological and reproductive healthy woman, cumulus cells could respond and adapt to oxidative stress-induced inflammatory and apoptotic signaling (*HMOX1* expression) during ovarian stimulation as reflected by the fact that its levels depend on the cohort oocyte’s maturation status. However, this pathway appears to be altered in AMA woman suggesting promissory biomarkers related to HMOX1 response. Additional functional studies are needed to deeply investigate this mechanism of adaptation to oxidative stress, as has also been indicated by Babayev and Duncan in their recent review where they propose a model of interaction between local oocyte microenvironment and oocyte quality [46].

Additionally, we suggest *VEGF* expression as a marker for fertilization rate, and that it may be a useful marker in oocyte preservation programs.

## Data Availability

All data produced in the present study are available upon reasonable request to the authors

## FIGURES

**Supplementary FIG 1**

**Relative expression levels of all selected genes among CCs**. Box-plot representation of gene expression (AU) of (a) aging-related genes and (b) Hypoxia-related genes. Red cross indicates outliers

## Statements and Declarations

## Funding

This work was supported by intramural funding of Clinica EUGIN and by the Secretary for Universities and Research of the Ministry of Economy and Knowledge of the Government of Catalonia (GENCAT 2015 DI 050) to D.C-B.

## Ethics approval

This study was performed in line with the principles of the Declaration of Helsinki.

Approval from the institutional Ethics Committee for Clinical Research of Clinica EUGIN was obtained before the beginning of this study (protocol code AGCC). All women were informed and consent to participate was collected in writing prior to their inclusion in the study.

## Competing interests

The authors have no competing interests to declare that are relevant to the content of this article.

## Conflict of interest

The authors declare that they have no conflict of interest.

## Acknowledgments

We would like to thank the laboratory technical staff from EUGIN for their help in sample handling.

## Authors contribution

M.B.: study design, data collection and analysis, manuscript preparation. D. C-B. and N.M.: data collection and analysis; R.V.: study implementation and supervision, expert knowledge, manuscript preparation.

## Notes

### Competing Interest Statement

The authors have declared no competing interest.

### Author Declarations

Ethics Committee for Clinical Research of Clinica EUGIN gave ethical approval forthis work

