## Supplementary figures and images for "The expression levels of *NOS2, HMOX1* and *VEGFC* in cumulus cells are markers of oocyte maturation and fertilization rate"

### Supplementery Figure 1

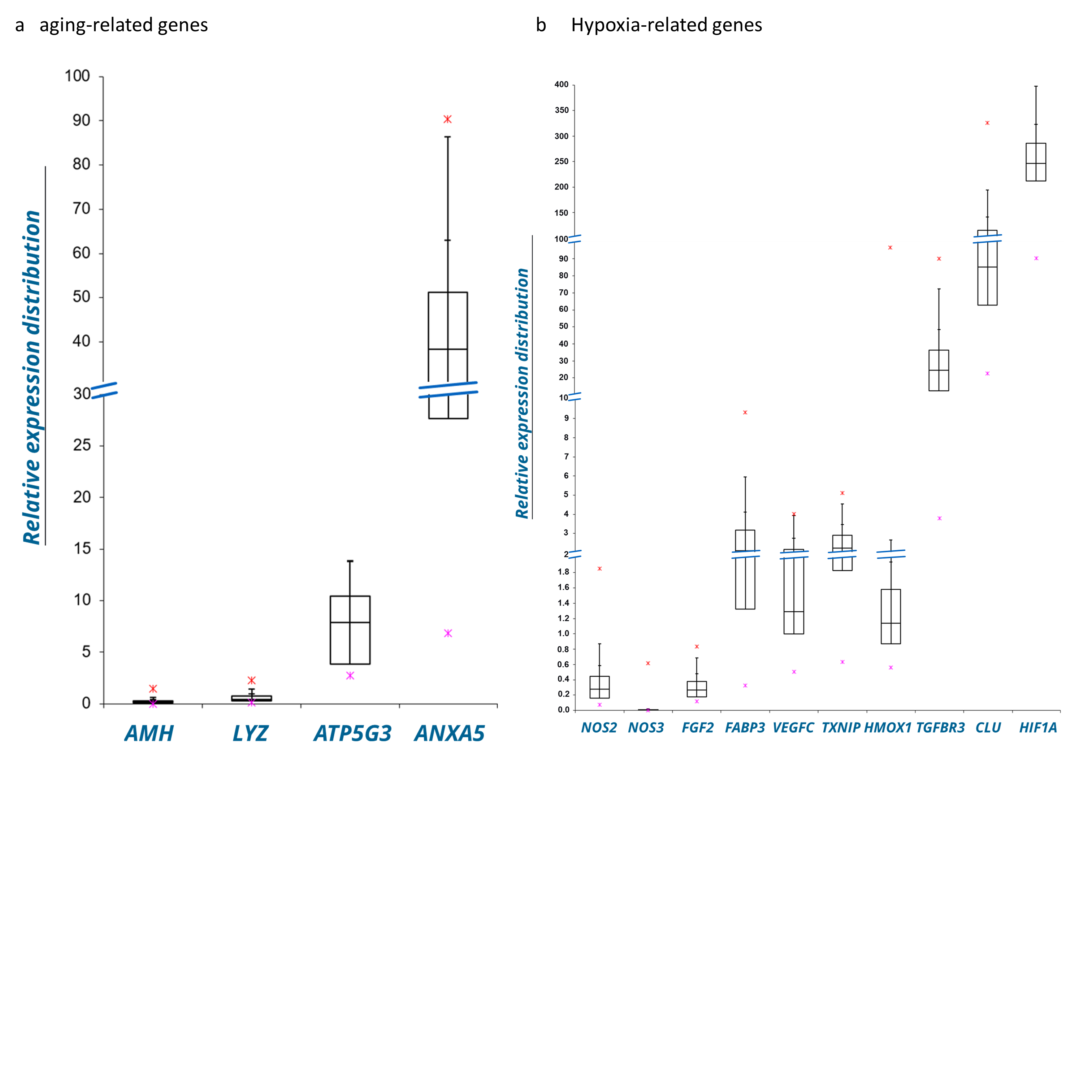
