## Supplementary Table 1 for "The expression levels of *NOS2, HMOX1* and *VEGFC* in cumulus cells are markers of oocyte maturation and fertilization rate"

**Supplementary Table 1.** Lists of 423 genes expressed in cumulus cells (independently of oocyte maturation) and annotated as aging-related genes in GenAge database.

| ***Gene symbol*** | **Gene name** | [**PANTHER Protein Class**](javascript:listAction(document.listForm,%20'list.do?filterLevel=1&sortField=PANTHER_PROTEIN_CLASS&listType=1&sortOrder=1&trackingId=D4F858B9CF193942790523089AB0E441&species=All%27)) |
| --- | --- | --- |
| *SIRT7* | NAD-dependent protein deacetylase sirtuin-7 | histone modifying enzyme (PC00261) |
| *BMI1* | Polycomb complex protein BMI-1 |  |
| *PPM1B* | Protein phosphatase 1B | protein phosphatase (PC00195) |
| *SIN3A* | Paired amphipathic helix protein Sin3a | chromatin/chromatin-binding, or -regulatory protein (PC00077) |
| *CSNK1A1* | Casein kinase I isoform alpha | non-receptor serine/threonine protein kinase (PC00167) |
| *ERCC8* | DNA excision repair protein ERCC-8 | DNA metabolism protein (PC00009) |
| *PIK3CB* | Phosphatidylinositol 4,5-bisphosphate 3-kinase catalytic subunit beta isoform | Kinase (PC00137) |
| *USP1* | Ubiquitin carboxyl-terminal hydrolase 1 | cysteine protease (PC00081) |
| *ZNF148* | Zinc finger protein 148 | C2H2 zinc finger transcription factor (PC00248) |
| *SRSF1* | Serine/arginine-rich splicing factor 1 | RNA splicing factor (PC00148) |
| *ESR1* | Estrogen receptor | C4 zinc finger nuclear receptor (PC00169) |
| *PCMT1* | Protein-L-isoaspartate(D-aspartate) O-methyltransferase | Methyltransferase (PC00155) |
| *VCP* | Transitional endoplasmic reticulum ATPase | primary active transporter (PC00068) |
| *PPARA* | Peroxisome proliferator-activated receptor alpha | C4 zinc finger nuclear receptor (PC00169) |
| *MAPT* | Microtubule-associated protein tau |  |
| *POT1* | Protection of telomeres protein 1 | DNA metabolism protein (PC00009) |
| *FGF2* | Fibroblast growth factor 2 | growth factor (PC00112) |
| *DHX9* | ATP-dependent RNA helicase A | RNA helicase (PC00032) |
| *TOP1* | DNA topoisomerase 1 | DNA topoisomerase (PC00017) |
| *WRN* | Werner syndrome ATP-dependent helicase | DNA helicase (PC00011) |
| *GPX4* | Phospholipid hydroperoxide glutathione peroxidase | Peroxidase (PC00180) |
| *POLA1* | DNA polymerase alpha catalytic subunit | DNA metabolism protein (PC00009) |
| *SMARCA4* | Transcription activator BRG1 | DNA helicase (PC00011) |
| *MSRA* | Mitochondrial peptide methionine sulfoxide reductase | Reductase (PC00198) |
| *TOP2A* | DNA topoisomerase 2-alpha | DNA metabolism protein (PC00009) |
| *AURKA* | Aurora kinase A | non-receptor serine/threonine protein kinase (PC00167) |
| *IRS2* | Insulin receptor substrate 2 |  |
| *RET* | Proto-oncogene tyrosine-protein kinase receptor Ret | transmembrane signal receptor (PC00197) |
| *NDRG1* | Protein NDRG1 | serine protease (PC00203) |
| *SOCS2* | Suppressor of cytokine signaling 2 | kinase modulator (PC00140) |
| *PTPN1* | Tyrosine-protein phosphatase non-receptor type 1 | protein phosphatase (PC00195) |
| *ING1* | Inhibitor of growth protein 1 | chromatin/chromatin-binding, or -regulatory protein (PC00077) |
| *BAX* | Apoptosis regulator BAX |  |
| *HSPA5* | Endoplasmic reticulum chaperone BiP | Hsp70 family chaperone (PC00027) |
| *YWHAZ* | 14-3-3 protein zeta/delta | scaffold/adaptor protein (PC00226) |
| *JAK2* | Tyrosine-protein kinase JAK2 | non-receptor tyrosine protein kinase (PC00168) |
| *NFE2L2* | Nuclear factor erythroid 2-related factor 2 | basic leucine zipper transcription factor (PC00056) |
| *PTEN* | Phosphatidylinositol 3,4,5-trisphosphate 3-phosphatase PTEN | protein phosphatase (PC00195) |
| *CDK4* | Cyclin-dependent kinase 4 | non-receptor serine/threonine protein kinase (PC00167) |
| *NFKB1* | Nuclear factor NF-kappa-B p105 subunit | Rel homology transcription factor (PC00252) |
| *ITPKB* | Inositol-trisphosphate 3-kinase B | Kinase (PC00137) |
| *EPS8* | Epidermal growth factor receptor kinase substrate 8 | scaffold/adaptor protein (PC00226) |
| *PRKCA* | Protein kinase C alpha type | non-receptor serine/threonine protein kinase (PC00167) |
| *MAP3K7* | Mitogen-activated protein kinase kinase kinase 7 | non-receptor serine/threonine protein kinase (PC00167) |
| *AXL* | Tyrosine-protein kinase receptor UFO | transmembrane signal receptor (PC00197) |
| *MAPKAPK5* | MAP kinase-activated protein kinase 5 | non-receptor serine/threonine protein kinase (PC00167) |
| *TXNIP* | Thioredoxin-interacting protein | ubiquitin-protein ligase (PC00234) |
| *HSP90AA1* | Heat shock protein HSP 90-alpha | Hsp90 family chaperone (PC00028) |
| *PIK3C2A* | Phosphatidylinositol 4-phosphate 3-kinase C2 domain-containing subunit alpha | Kinase (PC00137) |
| *ETS2* | Protein C-ets-2 | winged helix/forkhead transcription factor (PC00246) |
| *TGFBR3* | Transforming growth factor beta receptor type 3 | transmembrane signal receptor (PC00197) |
| *HTT* | Huntingtin |  |
| *PSEN1* | Presenilin-1 | aspartic protease (PC00053) |
| *NEK4* | Serine/threonine-protein kinase Nek4 | non-receptor serine/threonine protein kinase (PC00167) |
| *GHR* | Growth hormone receptor;GHR;ortholog | transmembrane signal receptor (PC00197) |
| *RAD21* | Double-strand-break repair protein rad21 homolog |  |
| *SUN1* | SUN domain-containing protein 1 | non-motor microtubule binding protein (PC00166) |
| *NEK6* | Serine/threonine-protein kinase Nek6 | non-receptor serine/threonine protein kinase (PC00167) |
| *PCGF2* | Polycomb group RING finger protein 2 |  |
| *FABP3* | Fatty acid-binding protein, heart | transfer/carrier protein (PC00219) |
| *EWSR1* | RNA-binding protein EWS | RNA metabolism protein (PC00031) |
| *CBX8* | Chromobox protein homolog 8 | chromatin/chromatin-binding, or -regulatory protein (PC00077) |
| *CSNK1E* | Casein kinase I isoform epsilon | non-receptor serine/threonine protein kinase (PC00167) |
| *EEF1A1* | Elongation factor 1-alpha 1 | translation factor (PC00223) |
| *NUDT1* | 7,8-dihydro-8-oxoguanine triphosphatase | Phosphatase (PC00181) |
| *NFKB2* | Nuclear factor NF-kappa-B p100 subunit | Rel homology transcription factor (PC00252) |
| *IL7R* | Interleukin-7 receptor subunit alpha | transmembrane signal receptor (PC00197) |
| *ANXA5* | Annexin A5 | calcium-binding protein (PC00060) |
| *PTPN11* | Tyrosine-protein phosphatase non-receptor type 11 | protein phosphatase (PC00195) |
| *RAE1* | mRNA export factor | RNA metabolism protein (PC00031) |
| *TPR* | Nucleoprotein TPR | primary active transporter (PC00068) |
| *CAV1* | Caveolin-1 | scaffold/adaptor protein (PC00226) |
| *MAGOH* | Protein mago nashi homolog |  |
| *SMARCB1* | SWI/SNF-related matrix-associated actin-dependent regulator of chromatin subfamily B member 1 | DNA metabolism protein (PC00009) |
| *SP1* | Transcription factor Sp1 | C2H2 zinc finger transcription factor (PC00248) |
| *UCP3* | Mitochondrial uncoupling protein 3 | secondary carrier transporter (PC00258) |
| *AGPAT2* | 1-acyl-sn-glycerol-3-phosphate acyltransferase beta | Acyltransferase (PC00042) |
| *NTN4* | Netrin-4 | extracellular matrix protein (PC00102) |
| *PPARG* | Peroxisome proliferator-activated receptor gamma | C4 zinc finger nuclear receptor (PC00169) |
| *VEGFC* | Vascular endothelial growth factor C | growth factor (PC00112) |
| *MT-CO1* | Cytochrome c oxidase subunit 1 | Oxidase (PC00175) |
| *SENP2* | Sentrin-specific protease 2 | Protease (PC00190) |
| *SENP7* | Sentrin-specific protease 7 | cysteine protease (PC00081) |
| *PTGS2* | Prostaglandin G/H synthase 2 | Oxigenase (PC00177) |
| *ERCC2* | General transcription and DNA repair factor IIH helicase subunit XPD | DNA helicase (PC00011) |
| *POLB* | DNA polymerase beta | DNA-directed DNA polymerase (PC00018) |
| *CKB* | Creatine kinase B-type | amino acid kinase (PC00045) |
| *EMD* | Emerin |  |
| *SIRT6* | NAD-dependent protein deacetylase sirtuin-6 | histone modifying enzyme (PC00261) |
| *SREBF1* | Sterol regulatory element-binding protein 1 |  |
| *ASF1A* | Histone chaperone ASF1A | chromatin/chromatin-binding, or -regulatory protein (PC00077) |
| *XPA* | DNA repair protein complementing XP-A cells | damaged DNA-binding protein (PC00086) |
| *PPM1D* | Protein phosphatase 1D | protein phosphatase (PC00195) |
| *CISD2* | CDGSH iron-sulfur domain-containing protein 2 |  |
| *NOTCH3* | Neurogenic locus notch homolog protein 3 |  |
| *TPP2* | Tripeptidyl-peptidase 2 | serine protease (PC00203) |
| *ARHGAP1* | Rho GTPase-activating protein 1 | GTPase-activating protein (PC00257) |
| *DUSP16* | Dual specificity protein phosphatase 16 | protein phosphatase (PC00195) |
| *HSPA1B* | Heat shock 70 kDa protein 1B | Hsp70 family chaperone (PC00027) |
| *FBXO31* | F-box only protein 31 | ubiquitin-protein ligase (PC00234) |
| *MAP2K3* | Dual specificity mitogen-activated protein kinase kinase 3 | non-receptor serine/threonine protein kinase (PC00167) |
| *MORC3* | MORC family CW-type zinc finger protein 3 |  |
| *ATF2* | Cyclic AMP-dependent transcription factor ATF-2 | basic leucine zipper transcription factor (PC00056) |
| *MAPK3* | Mitogen-activated protein kinase 3 | non-receptor serine/threonine protein kinase (PC00167) |
| *CDK7* | Cyclin-dependent kinase 7 | non-receptor serine/threonine protein kinase (PC00167) |
| *SENP1* | Sentrin-specific protease 1 | Protease (PC00190) |
| *DGCR8* | Microprocessor complex subunit DGCR8 | RNA processing factor (PC00147) |
| *CDKN1A* | Cyclin-dependent kinase inhibitor 1 | kinase inhibitor (PC00139) |
| *CEBPB* | CCAAT/enhancer-binding protein beta | basic leucine zipper transcription factor (PC00056) |
| *AKR1B1* | Aldo-keto reductase family 1 member B1 | Reductase (PC00198) |
| *STUB1* | E3 ubiquitin-protein ligase CHIP | ubiquitin-protein ligase (PC00234) |
| *FOXO3* | Forkhead box protein O3 | winged helix/forkhead transcription factor (PC00246) |
| *SIK1* | Serine/threonine-protein kinase SIK1 | non-receptor serine/threonine protein kinase (PC00167) |
| *GSTA4* | Glutathione S-transferase A4 | Transferase (PC00220) |
| *NOS2* | Nitric oxide synthase, inducible | Oxidoreductase (PC00176) |
| *BCL6* | B-cell lymphoma 6 protein | C2H2 zinc finger transcription factor (PC00248) |
| *CREB1* | Cyclic AMP-responsive element-binding protein 1 |  |
| *SQSTM1* | Sequestosome-1 |  |
| *HSF1* | Heat shock factor protein 1 | winged helix/forkhead transcription factor (PC00246) |
| *FOXO4* | Forkhead box protein O4 | winged helix/forkhead transcription factor (PC00246) |
| *GCLM* | Glutamate--cysteine ligase regulatory subunit | Ligase (PC00142) |
| *TRAP1* | Transforming growth factor-beta receptor-associated protein 1 |  |
| *CCNA2* | Cyclin-A2 | kinase activator (PC00138) |
| *TOP2B* | DNA topoisomerase 2-beta | DNA metabolism protein (PC00009) |
| *LGALS3* | Galectin-3 | extracellular matrix protein (PC00102) |
| *RAF1* | RAF proto-oncogene serine/threonine-protein kinase | non-receptor serine/threonine protein kinase (PC00167) |
| *MAPK9* | Mitogen-activated protein kinase 9 | non-receptor serine/threonine protein kinase (PC00167) |
| *PPP1CA* | Serine/threonine-protein phosphatase PP1-alpha catalytic subunit | protein phosphatase (PC00195) |
| *HBP1* | Glycosylphosphatidylinositol-anchored high-density lipoprotein-binding protein 1 | protein-binding activity modulator (PC00095) |
| *UBE2I* | SUMO-conjugating enzyme UBC9 | ubiquitin-protein ligase (PC00234) |
| *BLVRA* | Biliverdin reductase A | Dehydrogenase (PC00092) |
| *NFE2L1* | Endoplasmic reticulum membrane sensor NFE2L1 | basic leucine zipper transcription factor (PC00056) |
| *SOD2* | Superoxide dismutase [Mn], mitochondrial | Oxidoreductase (PC00176) |
| *LEO1* | RNA polymerase-associated protein LEO1 | DNA-directed RNA polymerase (PC00019) |
| *ING2* | Inhibitor of growth protein 2 | chromatin/chromatin-binding, or -regulatory protein (PC00077) |
| *HIVEP1* | Zinc finger protein 40 |  |
| *RPS6KA6* | Ribosomal protein S6 kinase alpha-6 | protein modifying enzyme (PC00260) |
| *CHEK2* | Serine/threonine-protein kinase Chk2 | non-receptor serine/threonine protein kinase (PC00167) |
| *FGFR1* | Fibroblast growth factor receptor 1 | transmembrane signal receptor (PC00197) |
| *MTOR* | Serine/threonine-protein kinase mTOR | non-receptor serine/threonine protein kinase (PC00167) |
| *PIK3CA* | Phosphatidylinositol 4,5-bisphosphate 3-kinase catalytic subunit alpha isoform | Kinase (PC00137) |
| *HSPA8* | Heat shock cognate 71 kDa protein | Hsp70 family chaperone (PC00027) |
| *HIF1A* | Hypoxia-inducible factor 1-alpha | basic helix-loop-helix transcription factor (PC00055) |
| *ID4* | DNA-binding protein inhibitor ID-4 | DNA-binding transcription factor (PC00218) |
| *MCL1* | Induced myeloid leukemia cell differentiation protein Mcl-1 |  |
| *NR3C1* | Glucocorticoid receptor | C4 zinc finger nuclear receptor (PC00169) |
| *TOP3B* | DNA topoisomerase 3-beta-1 | DNA metabolism protein (PC00009) |
| *SUPT5H* | Transcription elongation factor SPT5 |  |
| *SOX5* | Transcription factor SOX-5 |  |
| *PRMT6* | Protein arginine N-methyltransferase 6 | protein modifying enzyme (PC00260) |
| *TACC3* | Transforming acidic coiled-coil-containing protein 3 |  |
| *MLH1* | DNA mismatch repair protein Mlh1 | DNA metabolism protein (PC00009) |
| *BHLHE40* | Class E basic helix-loop-helix protein 40 | basic helix-loop-helix transcription factor (PC00055) |
| *NEK1* | Serine/threonine-protein kinase Nek1 | non-receptor serine/threonine protein kinase (PC00167) |
| *MT1E* | Metallothionein-1E | defense/immunity protein (PC00090) |
| *ACLY* | ATP-citrate synthase | Transferase (PC00220) |
| *RPA1* | DNA-directed RNA polymerase I subunit RPA1 | DNA-directed RNA polymerase (PC00019) |
| *BTG3* | Protein BTG3 |  |
| *MAP3K6* | Mitogen-activated protein kinase kinase kinase 6 | non-receptor serine/threonine protein kinase (PC00167) |
| *PAPPA* | Pappalysin-1 |  |
| *LATS1* | Serine/threonine-protein kinase LATS1 | non-receptor serine/threonine protein kinase (PC00167) |
| *DUSP3* | Dual specificity protein phosphatase 3 | protein phosphatase (PC00195) |
| *PDIK1L* | Serine/threonine-protein kinase PDIK1L | non-receptor serine/threonine protein kinase (PC00167) |
| *EPOR* | Erythropoietin receptor | transmembrane signal receptor (PC00197) |
| *STAT5A* | Signal transducer and activator of transcription 5A | DNA-binding transcription factor (PC00218) |
| *HSPA1A* | Heat shock 70 kDa protein 1A | Hsp70 family chaperone (PC00027) |
| *ATR* | Serine/threonine-protein kinase ATR | non-receptor serine/threonine protein kinase (PC00167) |
| *CLU* | Clusterin |  |
| *CENPA* | Histone H3-like centromeric protein A | chromatin/chromatin-binding, or -regulatory protein (PC00077) |
| *KDM5B* | Lysine-specific demethylase 5B | histone modifying enzyme (PC00261) |
| *EZH2* | Histone-lysine N-methyltransferase EZH2 | histone modifying enzyme (PC00261) |
| *BRD7* | Bromodomain-containing protein 7 | chromatin/chromatin-binding, or -regulatory protein (PC00077) |
| *SIRT3* | NAD-dependent protein deacetylase sirtuin-3, mitochondrial | histone modifying enzyme (PC00261) |
| *APP* | Amyloid-beta precursor protein | protease inhibitor (PC00191) |
| *SDHC* | Succinate dehydrogenase cytochrome b560 subunit, mitochondrial | Dehydrogenase (PC00092) |
| *HDAC4* | Histone deacetylase 4 | histone modifying enzyme (PC00261) |
| *TFDP1* | Transcription factor Dp-1 | DNA-binding transcription factor (PC00218) |
| *NCOR2* | Nuclear receptor corepressor 2 | chromatin/chromatin-binding, or -regulatory protein (PC00077) |
| *PDZD2* | PDZ domain-containing protein 2 | interleukin superfamily (PC00128) |
| *CREBBP* | CREB-binding protein | histone modifying enzyme (PC00261) |
| *PMVK* | Phosphomevalonate kinase | Kinase (PC00137) |
| *IKBKB* | Inhibitor of nuclear factor kappa-B kinase subunit beta | non-receptor serine/threonine protein kinase (PC00167) |
| *ERCC5* | DNA excision repair protein ERCC-5 | DNA metabolism protein (PC00009) |
| *HSPD1* | 60 kDa heat shock protein, mitochondrial |  |
| *PIK3R1* | Phosphatidylinositol 3-kinase regulatory subunit alpha | kinase modulator (PC00140) |
| *HMOX1* | Heme oxygenase 1 | Oxigenase (PC00177) |
| *XRCC6* | X-ray repair cross-complementing protein 6 | DNA helicase (PC00011) |
| *GSK3A* | Glycogen synthase kinase-3 alpha | non-receptor serine/threonine protein kinase (PC00167) |
| *IRF3* | Interferon regulatory factor 3 | winged helix/forkhead transcription factor (PC00246) |
| *SOCS1* | Suppressor of cytokine signaling 1 | kinase modulator (PC00140) |
| *SORBS2* | Sorbin and SH3 domain-containing protein 2 | scaffold/adaptor protein (PC00226) |
| *COQ7* | 5-demethoxyubiquinone hydroxylase, mitochondrial |  |
| *CDK6* | Cyclin-dependent kinase 6 | non-receptor serine/threonine protein kinase (PC00167) |
| *WT1* | Wilms tumor protein | C2H2 zinc finger transcription factor (PC00248) |
| *UCHL1* | Ubiquitin carboxyl-terminal hydrolase isozyme L1 | cysteine protease (PC00081) |
| *VEGFA* | Vascular endothelial growth factor A | growth factor (PC00112) |
| *MXD4* | Max dimerization protein 4 | basic helix-loop-helix transcription factor (PC00055) |
| *ZMPSTE24* | CAAX prenyl protease 1 homolog | Metalloprotease (PC00153) |
| *SIN3B* | Paired amphipathic helix protein Sin3b | chromatin/chromatin-binding, or -regulatory protein (PC00077) |
| *MXI1* | Max-interacting protein 1 | basic helix-loop-helix transcription factor (PC00055) |
| *GATA4* | Transcription factor GATA-4 | DNA-binding transcription factor (PC00218) |
| *LIMK1* | LIM domain kinase 1 |  |
| *NADK* | NAD kinase | nucleotide kinase (PC00172) |
| *DGAT1* | Diacylglycerol O-acyltransferase 1 | Acyltransferase (PC00042) |
| *PRKCD* | Protein kinase C delta type | non-receptor serine/threonine protein kinase (PC00167) |
| *IGF2* | Insulin-like growth factor II | growth factor (PC00112) |
| *ERCC6* | DNA excision repair protein ERCC-6 | damaged DNA-binding protein (PC00086) |
| *GSS* | Glutathione synthetase | Ligase (PC00142) |
| *UBB* | Polyubiquitin-B |  |
| *MAP2K7* | Dual specificity mitogen-activated protein kinase kinase 7 | non-receptor serine/threonine protein kinase (PC00167) |
| *IGF1R* | Insulin-like growth factor 1 receptor | transmembrane signal receptor (PC00197) |
| *FAS* | Tumor necrosis factor receptor superfamily member 6 | transmembrane signal receptor (PC00197) |
| *CDK1* | Cyclin-dependent kinase 1 | non-receptor serine/threonine protein kinase (PC00167) |
| *APOE* | Apolipoprotein E | Apolipoprotein (PC00052) |
| *FEN1* | Flap endonuclease 1 | Exodeoxyribonuclease (PC00098) |
| *STAT3* | Signal transducer and activator of transcription 3 | DNA-binding transcription factor (PC00218) |
| *PBRM1* | Protein polybromo-1 | chromatin/chromatin-binding, or -regulatory protein (PC00077) |
| *ITPK1* | Inositol-tetrakisphosphate 1-kinase | Kinase (PC00137) |
| *MVK* | Mevalonate kinase | carbohydrate kinase (PC00065) |
| *MXD1* | Max dimerization protein 1 | basic helix-loop-helix transcription factor (PC00055) |
| *ATP5O* | ATP synthase subunit O, mitochondrial | ATP synthase (PC00002) |
| *ATP5G3* | ATP synthase F(0) complex subunit C3, mitochondrial | ATP synthase (PC00002) |
| *HDAC1* | Histone deacetylase 1 | histone modifying enzyme (PC00261) |
| *MDM2* | E3 ubiquitin-protein ligase Mdm2 |  |
| *SHC1* | SHC-transforming protein 1 | scaffold/adaptor protein (PC00226) |
| *SRC* | Proto-oncogene tyrosine-protein kinase Src | non-receptor tyrosine protein kinase (PC00168) |
| *TP53BP1* | TP53-binding protein 1 | DNA metabolism protein (PC00009) |
| *ABL1* | Tyrosine-protein kinase ABL1 | non-receptor tyrosine protein kinase (PC00168) |
| *PML* | Protein PML | ubiquitin-protein ligase (PC00234) |
| *GPX1* | Glutathione peroxidase 1 | Peroxidase (PC00180) |
| *NBN* | Nibrin | damaged DNA-binding protein (PC00086) |
| *SMG1* | Serine/threonine-protein kinase SMG1 | non-receptor serine/threonine protein kinase (PC00167) |
| *HIC1* | Hypermethylated in cancer 1 protein | C2H2 zinc finger transcription factor (PC00248) |
| *PDGFRA* | Platelet-derived growth factor receptor alpha | transmembrane signal receptor (PC00197) |
| *SOD1* | Superoxide dismutase [Cu-Zn] | Oxidoreductase (PC00176) |
| *MAGOHB* | Protein mago nashi homolog 2 |  |
| *TRPV1* | Transient receptor potential cation channel subfamily V member 1 | ion channel (PC00133) |
| *BRCA1* | Breast cancer type 1 susceptibility protein | ubiquitin-protein ligase (PC00234) |
| *ENDOG* | Endonuclease G, mitochondrial |  |
| *PDGFRB* | Platelet-derived growth factor receptor beta | transmembrane signal receptor (PC00197) |
| *GLB1* | Beta-galactosidase | Galactosidase (PC00104) |
| *ITSN2* | Intersectin-2 | membrane traffic protein (PC00150) |
| *STK11* | Serine/threonine-protein kinase STK11 |  |
| *CDC42* | Cell division control protein 42 homolog | small GTPase (PC00208) |
| *INSR* | Insulin receptor | transmembrane signal receptor (PC00197) |
| *G6PD* | Glucose-6-phosphate 1-dehydrogenase | Dehydrogenase (PC00092) |
| *TLR3* | Toll-like receptor 3 | transmembrane signal receptor (PC00197) |
| *FLT1* | Vascular endothelial growth factor receptor 1 | transmembrane signal receptor (PC00197) |
| *TERF2* | Telomeric repeat-binding factor 2 |  |
| *EEF1E1* | Eukaryotic translation elongation factor 1 epsilon-1 | translation elongation factor (PC00222) |
| *HRAS* | GTPase HRas | small GTPase (PC00208) |
| *ID1* | DNA-binding protein inhibitor ID-1 | DNA-binding transcription factor (PC00218) |
| *PSMD14* | 26S proteasome non-ATPase regulatory subunit 14 | translation initiation factor (PC00224) |
| *MCRS1* | Microspherule protein 1 |  |
| *SPIN1* | Spindlin-1 | chromatin/chromatin-binding, or -regulatory protein (PC00077) |
| *TERF1* | Telomeric repeat-binding factor 1 |  |
| *NOS3* | Nitric oxide synthase, endothelial | Oxidoreductase (PC00176) |
| *TGFB1I1* | Transforming growth factor beta-1-induced transcript 1 protein | actin or actin-binding cytoskeletal protein (PC00041) |
| *DDIT3* | DNA damage-inducible transcript 3 protein |  |
| *POLD1* | DNA polymerase delta catalytic subunit | DNA metabolism protein (PC00009) |
| *MAPK14* | Mitogen-activated protein kinase 14 | non-receptor serine/threonine protein kinase (PC00167) |
| *EP300* | Histone acetyltransferase p300 | histone modifying enzyme (PC00261) |
| *S100B* | Protein S100-B | calmodulin-related (PC00061) |
| *PARP1* | Poly [ADP-ribose] polymerase 1 | DNA metabolism protein (PC00009) |
| *PATZ1* | POZ-, AT hook-, and zinc finger-containing protein 1 | C2H2 zinc finger transcription factor (PC00248) |
| *JUN* | Transcription factor AP-1 | basic leucine zipper transcription factor (PC00056) |
| *TCF3* | Transcription factor E2-alpha | basic helix-loop-helix transcription factor (PC00055) |
| *PDCD10* | Programmed cell death protein 10 |  |
| *APEX1* | DNA-(apurinic or apyrimidinic site) endonuclease |  |
| *LMNB1* | Lamin-B1 |  |
| *MYLK* | Myosin light chain kinase, smooth muscle |  |
| *DHCR24* | Delta(24)-sterol reductase | Reductase (PC00198) |
| *PEBP1* | Phosphatidylethanolamine-binding protein 1 | protease inhibitor (PC00191) |
| *PSMB5* | Proteasome subunit beta type-5 | Protease (PC00190) |
| *BLM* | Bloom syndrome protein | DNA helicase (PC00011) |
| *IGFBP2* | Insulin-like growth factor-binding protein 2 | protease inhibitor (PC00191) |
| *CDKN2AIP* | CDKN2A-interacting protein | DNA-binding transcription factor (PC00218) |
| *ERCC4* | DNA repair endonuclease XPF | Endodeoxyribonuclease (PC00093) |
| *BUB3* | Mitotic checkpoint protein BUB3 | RNA metabolism protein (PC00031) |
| *RUVBL2* | RuvB-like 2 |  |
| *EFEMP1* | EGF-containing fibulin-like extracellular matrix protein 1 | extracellular matrix structural protein (PC00103) |
| *ATF7IP* | Activating transcription factor 7-interacting protein 1 | transcription cofactor (PC00217) |
| *SUMO1* | Small ubiquitin-related modifier 1 |  |
| *ZFP36* | mRNA decay activator protein ZFP36 | RNA metabolism protein (PC00031) |
| *YAP1* | Transcriptional coactivator YAP1 | transcription cofactor (PC00217) |
| *PRKCH* | Protein kinase C eta type | non-receptor serine/threonine protein kinase (PC00167) |
| *GNG11* | Guanine nucleotide-binding protein G(I)/G(S)/G(O) subunit gamma-11 | heterotrimeric G-protein (PC00117) |
| *CLOCK* | Circadian locomoter output cycles protein kaput |  |
| *RECQL4* | ATP-dependent DNA helicase Q4 | DNA helicase (PC00011) |
| *AKT1* | RAC-alpha serine/threonine-protein kinase | non-receptor serine/threonine protein kinase (PC00167) |
| *ZMAT3* | Zinc finger matrin-type protein 3 | RNA processing factor (PC00147) |
| *GCLC* | Glutamate--cysteine ligase catalytic subunit | Ligase (PC00142) |
| *EEF2* | Elongation factor 2 | translation elongation factor (PC00222) |
| *HSPA9* | Stress-70 protein, mitochondrial | Hsp70 family chaperone (PC00027) |
| *YPEL3* | Protein yippee-like 3 | ubiquitin-protein ligase (PC00234) |
| *LYZ* | Lysozyme C | Glycosidase (PC00110) |
| *CSNK2A1* | Casein kinase II subunit alpha |  |
| *RBX1* | E3 ubiquitin-protein ligase RBX1 | ubiquitin-protein ligase (PC00234) |
| *PRDX1* | Peroxiredoxin-1 | Peroxidase (PC00180) |
| *WWP1* | NEDD4-like E3 ubiquitin-protein ligase WWP1 | ubiquitin-protein ligase (PC00234) |
| *AAK1* | AP2-associated protein kinase 1 |  |
| *CHEK1* | Serine/threonine-protein kinase Chk1 | non-receptor serine/threonine protein kinase (PC00167) |
| *STAT5B* | Signal transducer and activator of transcription 5B | DNA-binding transcription factor (PC00218) |
| *PRKDC* | DNA-dependent protein kinase catalytic subunit | non-receptor serine/threonine protein kinase (PC00167) |
| *TP53* | Cellular tumor antigen p53 | P53-like transcription factor (PC00253) |
| *TMSB4X* | Thymosin beta-4 | actin or actin-binding cytoskeletal protein (PC00041) |
| *FXR1* | Fragile X mental retardation syndrome-related protein 1 | translation factor (PC00223) |
| *TRIM28* | Transcription intermediary factor 1-beta | ubiquitin-protein ligase (PC00234) |
| *GDF11* | Growth/differentiation factor 11 | growth factor (PC00112) |
| *MAPK8* | Mitogen-activated protein kinase 8 | non-receptor serine/threonine protein kinase (PC00167) |
| *PYCR1* | Pyrroline-5-carboxylate reductase 1, mitochondrial | Reductase (PC00198) |
| *TXN* | Thioredoxin | Oxidoreductase (PC00176) |
| *MAP2K2* | Dual specificity mitogen-activated protein kinase kinase 2 | non-receptor serine/threonine protein kinase (PC00167) |
| *BSCL2* | Seipin |  |
| *NFKBIA* | NF-kappa-B inhibitor alpha |  |
| *A2M* | Alpha-2-macroglobulin | protease inhibitor (PC00191) |
| *CXCL1* | Growth-regulated alpha protein | Chemokine (PC00074) |
| *AR* | Androgen receptor | C4 zinc finger nuclear receptor (PC00169) |
| *PAK4* | Serine/threonine-protein kinase PAK 4 | non-receptor serine/threonine protein kinase (PC00167) |
| *PIAS4* | E3 SUMO-protein ligase PIAS4 | ubiquitin-protein ligase (PC00234) |
| *BCL2* | Apoptosis regulator Bcl-2 |  |
| *LEPR* | Leptin receptor | transmembrane signal receptor (PC00197) |
| *IGFBP1* | Insulin-like growth factor-binding protein 1 | protease inhibitor (PC00191) |
| *IGFBP3* | Insulin-like growth factor-binding protein 3 | protease inhibitor (PC00191) |
| *PIN1* | Peptidyl-prolyl cis-trans isomerase NIMA-interacting 1 | Chaperone (PC00072) |
| *HMGB2* | High mobility group protein B2 | chromatin/chromatin-binding, or -regulatory protein (PC00077) |
| *CBX7* | Chromobox protein homolog 7 |  |
| *CDKN1C* | Cyclin-dependent kinase inhibitor 1C | kinase inhibitor( PC00139) |
| *XRCC5* | X-ray repair cross-complementing protein 5 | DNA helicase (PC00011) |
| *POLG* | DNA polymerase subunit gamma-1 | DNA-directed DNA polymerase (PC00018) |
| *ASPH* | Aspartyl/asparaginyl beta-hydroxylase | protein modifying enzyme (PC00260) |
| *GAPDH* | Glyceraldehyde-3-phosphate dehydrogenase | Dehydrogenase (PC00092) |
| *RICTOR* | Rapamycin-insensitive companion of mTOR |  |
| *SMURF2* | E3 ubiquitin-protein ligase SMURF2 | ubiquitin-protein ligase (PC00234) |
| *FOXO1* | Forkhead box protein O1 | winged helix/forkhead transcription factor (PC00246) |
| *SERPINE1* | Plasminogen activator inhibitor 1 | protease inhibitor (PC00191) |
| *RAD52* | DNA repair protein RAD52 homolog | DNA metabolism protein (PC00009) |
| *BRAF* | Serine/threonine-protein kinase B-raf | non-receptor serine/threonine protein kinase (PC00167) |
| *ETS1* | Protein C-ets-1 | winged helix/forkhead transcription factor (PC00246) |
| *NUAK1* | NUAK family SNF1-like kinase 1 | non-receptor serine/threonine protein kinase (PC00167) |
| *BAG3* | BAG family molecular chaperone regulator 3 | Chaperone (PC00072) |
| *ERCC1* | DNA excision repair protein ERCC-1 | Endodeoxyribonuclease (PC00093) |
| *CAT* | Catalase | Peroxidase (PC00180) |
| *STK40* | Serine/threonine-protein kinase 40 | non-receptor serine/threonine protein kinase (PC00167) |
| *GRB2* | Growth factor receptor-bound protein 2 |  |
| *KDM4A* | Lysine-specific demethylase 4A | histone modifying enzyme (PC00261) |
| *FASTK* | Fas-activated serine/threonine kinase | RNA processing factor (PC00147) |
| *IRS1* | Insulin receptor substrate 1 |  |
| *RELA* | Transcription factor p65 | Rel homology transcription factor (PC00252) |
| *ERCC3* | General transcription and DNA repair factor IIH helicase subunit XPB | DNA helicase (PC00011) |
| *HOXC4* | Homeobox protein Hox-C4 |  |
| *GSR* | Glutathione reductase, mitochondrial | Reductase (PC00198) |
| *EIF5A2* | Eukaryotic translation initiation factor 5A-2 | translation initiation factor (PC00224) |
| *LMNA* | Prelamin-A/C |  |
| *NINJ1* | Ninjurin-1 | cell adhesion molecule (PC00069) |
| *MYC* | Myc proto-oncogene protein | basic helix-loop-helix transcription factor (PC00055) |
| *PCNA* | Proliferating cell nuclear antigen | DNA polymerase processivity factor (PC00015) |
| *RNASEL* | 2-5A-dependent ribonuclease | Endoribonuclease (PC00094) |
| *AIFM1* | Apoptosis-inducing factor 1, mitochondrial | Oxidoreductase (PC00176) |
| *SGK1* | Serine/threonine-protein kinase Sgk1 | non-receptor serine/threonine protein kinase (PC00167) |
| *FOXM1* | Forkhead box protein M1 |  |
| *EGFR* | Epidermal growth factor receptor | transmembrane signal receptor (PC00197) |
| *JUND* | Transcription factor jun-D | basic leucine zipper transcription factor (PC00056) |
| *STK32C* | Serine/threonine-protein kinase 32C | non-receptor serine/threonine protein kinase (PC00167) |
| *ADCK5* | Uncharacterized aarF domain-containing protein kinase 5 | ATP-binding cassette (ABC) transporter (PC00003) |
| *FOS* | Proto-oncogene c-Fos | basic leucine zipper transcription factor (PC00056) |
| *MED1* | Mediator of RNA polymerase II transcription subunit 1 | general transcription factor (PC00259) |
| *TFAP4* | Transcription factor AP-4 | DNA-binding transcription factor (PC00218) |
| *BAK1* | Bcl-2 homologous antagonist/killer |  |
| *LIMA1* | LIM domain and actin-binding protein 1 | actin or actin-binding cytoskeletal protein (PC00041) |
| *ARNTL* | Aryl hydrocarbon receptor nuclear translocator-like protein 1 | basic helix-loop-helix transcription factor (PC00055) |
| *SLC16A7* | Monocarboxylate transporter 2 | Transporter (PC00227) |
| *TYK2* | Non-receptor tyrosine-protein kinase TYK2 | non-receptor tyrosine protein kinase (PC00168) |
| *MAP2K1* | Dual specificity mitogen-activated protein kinase kinase 1 | non-receptor serine/threonine protein kinase (PC00167) |
| *CDKN1B* | Cyclin-dependent kinase inhibitor 1B | kinase inhibitor (PC00139) |
| *DEK* | Protein DEK | chromatin/chromatin-binding, or -regulatory protein (PC00077) |
| *PTK2* | Focal adhesion kinase 1 | non-receptor tyrosine protein kinase (PC00168) |
| *UCP2* | Mitochondrial uncoupling protein 2 | secondary carrier transporter (PC00258) |
| *RUNX1* | Runt-related transcription factor 1 | Runt transcription factor (PC00254) |
| *MECP2* | Methyl-CpG-binding protein 2 | chromatin/chromatin-binding, or -regulatory protein (PC00077) |
| *IRF7* | Interferon regulatory factor 7 | winged helix/forkhead transcription factor (PC00246) |
| *DDB2* | DNA damage-binding protein 2 | damaged DNA-binding protein (PC00086) |
| *MDH1* | Malate dehydrogenase, cytoplasmic | Dehydrogenase (PC00092) |
| *SIRT1* | NAD-dependent protein deacetylase sirtuin-1 | histone modifying enzyme (PC00261) |
| *TAF1* | Transcription initiation factor TFIID subunit 1 | general transcription factor (PC00259) |
| *ARPC1B* | Actin-related protein 2/3 complex subunit 1B | actin or actin-binding cytoskeletal protein (PC00041) |
| *E2F1* | Transcription factor E2F1 | general transcription factor (PC00259) |
| *MAP3K5* | Mitogen-activated protein kinase kinase kinase 5 | non-receptor serine/threonine protein kinase (PC00167) |
| *ATM* | Serine-protein kinase ATM | non-receptor serine/threonine protein kinase (PC00167) |
| *RSL1D1* | Ribosomal L1 domain-containing protein 1 | ribosomal protein (PC00202) |
| *SLC13A3* | Solute carrier family 13 member 3 | secondary carrier transporter (PC00258) |
| *GSK3B* | Glycogen synthase kinase-3 beta | non-receptor serine/threonine protein kinase (PC00167) |
| *CDK2AP1* | Cyclin-dependent kinase 2-associated protein 1 | kinase inhibitor (PC00139) |
| *TGFB1* | Transforming growth factor beta-1 proprotein | growth factor (PC00112) |
| *PEX5* | Peroxisomal targeting signal 1 receptor | membrane trafficking regulatory protein (PC00151) |
| *HTRA2* | Serine protease HTRA2, mitochondrial | serine protease (PC00203) |
| *ERRFI1* | ERBB receptor feedback inhibitor 1 |  |
| *GRK6* | G protein-coupled receptor kinase 6 | non-receptor serine/threonine protein kinase (PC00167) |
| *CDK18* | Cyclin-dependent kinase 18 | non-receptor serine/threonine protein kinase (PC00167) |
| *PDPK1* | 3-phosphoinositide-dependent protein kinase 1 | non-receptor serine/threonine protein kinase (PC00167) |
| *PCK1* | Phosphoenolpyruvate carboxykinase, cytosolic [GTP] | Kinase (PC00137) |
| *ERBB2* | Receptor tyrosine-protein kinase erbB-2 | transmembrane signal receptor (PC00197) |
| *XAF1* | XIAP-associated factor 1 |  |
| *HDAC3* | Histone deacetylase 3 | histone modifying enzyme (PC00261) |
| *APTX* | Aprataxin | damaged DNA-binding protein (PC00086) |
| *PEX19* | Peroxisomal biogenesis factor 19 | membrane traffic protein (PC00150) |
| *IGFBP5* | Insulin-like growth factor-binding protein 5 | protease inhibitor (PC00191) |
| *PROX1* | Prospero homeobox protein 1 | homeodomain transcription factor (PC00119) |
| *PRPF19* | Pre-mRNA-processing factor 19 | RNA processing factor (PC00147) |
| *RGN* | Regucalcin | Esterase (PC00097) |
| *HDAC2* | Histone deacetylase 2 | histone modifying enzyme (PC00261) |
| *HMGB1* | High mobility group protein B1 | chromatin/chromatin-binding, or -regulatory protein (PC00077) |
| *GSTP1* | Glutathione S-transferase P | Transferase (PC00220) |
| *NCOR1* | Nuclear receptor corepressor 1 | chromatin/chromatin-binding, or -regulatory protein (PC00077) |
| *PIM1* | Serine/threonine-protein kinase pim-1 | non-receptor serine/threonine protein kinase (PC00167) |
| *HJURP* | Holliday junction recognition protein |  |
| *BDNF* | Brain-derived neurotrophic factor | neurotrophic factor (PC00163) |
| *PTTG1* | Securin |  |
| *PNPT1* | Polyribonucleotide nucleotidyltransferase 1, mitochondrial | Nucleotidyltransferase (PC00174) |
| *GRN* | Progranulin |  |
| *EGR1* | Early growth response protein 1 | C2H2 zinc finger transcription factor (PC00248) |
| *SPOP* | Speckle-type POZ protein | ubiquitin-protein ligase (PC00234) |
| *TBP* | TATA-box-binding protein | general transcription factor (PC00259) |
