## Supplementary Table 2 for "The expression levels of *NOS2, HMOX1* and *VEGFC* in cumulus cells are markers of oocyte maturation and fertilization rate"

**Supplementary Table 2.** Primer Sequences Used for Real-Time qPCR analysis

|  | **Gene Symbol** | **Gene name** | **Ensembl accession no.** | **Primer sequence F (5’→3’)** | **Primer sequence R (5’→3’)** | **Efficiency (%)** |
| --- | --- | --- | --- | --- | --- | --- |
| **Reference genes**  **(HK)** | *ACTB* | Actin beta | ENSG00000075624 | GGACTTCGAGCAAGAGATGG | AGCACTGTGTTGGCGTACAG | 74.1 |
|  | *GAPDH* | Glyceraldehyde-3-phosphate dehydrogenase | ENSG00000111640 | GAGTCAACGGATTTGGTCGT | TTGATTTTGGAGGGATCTCG | 83.4 |
|  | *GUSB* | Glucuronidase beta | ENSG00000169919 | AAACGATTGCAGGGTTTCAC | CTCTCGTCGGTGACTGTTCA | 79.0 |
|  | *RPLP0* | 60S acidic ribosomal protein P0 | ENSG00000089157 | GGCGACCTGGAAGTCCAACT | CCATCAGCACCACAGCCTTC | 82.3 |
|  | *SDHA* | Succinate dehydrogenase flavoprotein subunit A | ENSG00000073578 | TGGGAACAAGAGGGCATCTG | CCACCACTGCATCAAATTCATG | 79.7 |
|  | *TBP* | TATA-box binding protein | ENSG00000112592 | TATAATCCCAAGCGGTTTGC | GCTGGAAAACCCAACTTCTG | 92.8 |
|  | *UBC* | Ubiquitin C | ENSG00000150991 | ATTTGGGTCGCGGTTCTTG | TGCCTTGACATTCTCGATGGT | 97.6 |
|  | *YWHAZ* | 14-3-3 protein zeta/delta | ENSG00000164924 | ACTTTTGGTACATTGTGGCTTCAA | CCGCCAGGACAAACCAGTAT | 86.2 |
|  | *18S* | 18S ribosomal pseudogene | ENST00000445125 | TCTGTCGAGATCACAAGTTGC | AGCATAGAAGATGATACCCGTGT | 86.8 |
| **Aging-related**  **genes** | *AMH* | anti-Mullerian hormone | ENSG00000104899 | GCTGCCTTGCCCTCTCTAC | GAACCTCAGCGAGGGTGTT | 72.2 |
|  | *ANXA5* | Annexin A5 | ENSG00000164111 | GACCCTCTATTATGCTATGAAG | TCCTAAACTCCTTCCTGATG | 86.9 |
|  | *ATP5G3* | ATP synthase F(0) complex subunit C3 | ENSG00000249253 | ACGTCGCCTGTCACCCAATA | TGGTCGAGATAACACTGATGCAGA | 79.5 |
|  | *FGF2* | Fibroblast growth factor 2 | ENSG00000138685 | CCGTTACCTGGCTATGAAGG | ACTGCCCAGTTCGTTTCAGT | 80.4 |
|  | *LYZ* | Lysozyme | ENSG00000090382 | CCGCTACTGGTGTAATGATGG | CATCAGCGATGTTATCTTGCAG | 95.2 |
| **Hypoxia-related**  **genes** | *CLU* | Clusterin | ENSG00000120885 | GCAAGACACTGCTCAGCAAC | TCAGGCAGGGCTTACACTCT | 90 |
|  | *FABP3* | Fatty acid binding protein 3 | ENSG00000121769 | GGTGGAGTTCGATGAGACAA | TCAATTAGCTCCCGCACAAG | 90 |
|  | *HMOX1* | Heme oxygenase 1 | ENSG00000100292 | AGGAGGAGATTGAGCGCCAC | GCTTCACATAGCGCTGCATG | 91.9 |
|  | *HIF1A* | Hypoxia inducible factor 1 subunit alpha | ENSG00000100644 | TTCACCTGAGCCTAATAGTCC | CAAGTCTAAATCTGTGTCCTG | 110.4 |
|  | *NOS2* | Nitric oxide synthase 2 | ENSG00000007171 | AGGAGGAGATGCTGGAGATG | ACATCCCCGCAAACATAGAG | 106.7 |
|  | *NOS3* | Nitric oxide synthase 3 | ENSG00000164867 | ACCCTCACCGCTACAACATC | CTGGCCTTCTGCTCATTCTC | 90.7 |
|  | *TGFBR3* | Transforming growth factor beta receptor 3 | ENSG00000069702 | TGGAGTCTCCTCTGAATGGCTG | CCATTATCACCTGACTCCAGATC | 84.7 |
|  | *TXNIP* | Thioredoxin interacting protein | ENSG00000265972 | GCAAGCCTAATGGCTACTCG | TTGAAGGATGTTCCCAGAGG | 92.8 |
|  | *VEGFC* | vascular endothelial growth factor C | ENSG00000150630 | GGATGCTGGAGATGACTCAA | TTCATCCAGCTCCTTGTTTG | 83.9 |
